# Self-Collected Finger-Prick Blood for Gene Expression Profiling: Unveiling Early Immune Responses in Mild COVID-19

**DOI:** 10.1101/2025.05.06.25327063

**Authors:** Rodolphe Thiébaut, Edouard Lhomme, Hakim Hocini, Isabelle Pellegrin, Andrea Boizard-Moracchini, Alexandre Duvignaud, Maud Perpère, Mélanie Huchon, Mélanie Prague, Christine Lacabaratz, Mathieu Surenaud, Xavier Anglaret, Denis Malvy, Boris Hejblum, COVERAGE study group, Yves Levy

## Abstract

**Background:** Whole blood gene expression analysis is essential for understanding molecular host responses and identifying markers of disease severity. Self-collected finger-prick capillary blood provides a promising alternative to venous sampling, yet its application to transcriptomics is still underexplored.

**Methods:** COVERAGE-Immuno sub-study is an ancillary study of the COVERAGE France platform trial (NCT04356495), a randomized controlled trial of COVID-19 early treatment in at risk patients with mild COVID-19 monitored at home. Participants recruited additional sampling for in-depth, repeated evaluation of the immunological markers and gene expression data. We compared gene expression data obtained simultaneously from venous blood (Tempus tube) and ultralow-volume fingerstick samples.

**Findings:** Our analysis revealed excellent concordance between the two sampling methods at the gene level (ICC=0·74) and a nearly perfect agreement at the geneset level. Gene expression profiles correlated well with deconvoluted cell frequencies of B and CD8+ T cells and serum biomarker IL-1b and IP-10 dynamics, offering valuable insights into SARS-CoV-2 pathophysiology. Hence, the dynamics of the immune response could be analyzed at a daily resolution, confirming early signals of neutrophil activation, interferon signaling, erythroid cell involvement, and inflammation pathways during the early stages of mild COVID-19.

**Interpretation:** These findings validate the feasibility and reliability of fingerstick sampling for at-home use, providing a unique tool for advanced clinical research and precision medicine.

**Funding:** EIT Health (Grant number: 20874 COVERAGE-Immuno)

**Research in context:** *Evidence before this study:* Gene expression analysis of whole blood is widely used to investigate host immune responses and identify biomarkers of disease severity. While venous blood is the standard for transcriptomic studies, capillary blood sampling via self-collected finger-prick offers a less invasive and more accessible alternative, particularly for remote monitoring. However, its application to transcriptomics remains underexplored. Prior studies have demonstrated the feasibility of capillary blood for certain molecular analyses, but its reliability compared to venous sampling for gene expression profiling is not well established.

*Added value of this study:* This study provides the first comprehensive comparison of transcriptomic profiles from venous and capillary blood in a clinical trial setting. We show that gene expression measurements from self-collected fingerstick samples exhibit strong concordance with venous samples (ICC = 0·74) and near-perfect agreement at the gene set level. These findings demonstrate that capillary sampling can capture dynamic immune responses, including neutrophil activation, interferon signaling, and inflammation pathways, with correlations to key immune biomarkers (IL-1b, IP-10).

*Implications of all the available evidence:* Fingerstick-based gene expression analysis is a reliable and scalable alternative to venous blood sampling, offering a practical solution for at-home immune monitoring. These findings support its integration into clinical research and precision medicine approaches, enabling frequent, minimally invasive monitoring of immune responses in infectious diseases and beyond.

## Introduction

At the era of systems biology and targeted therapies, analysis of whole blood gene changes provides key information about molecular host responses to diseases, infections or therapeutic interventions and possibly markers or correlates of disease severity^1,2^. However, a full use of these approaches requires repeated sampling, especially around interventions such as vaccine injections or disease onset. Due to its practical constraints, venous blood sampling is not feasible at a high frequency and by the participant him/herself. This issue becomes even more pronounced when considering certain groups as healthy volunteers, children, or in some settings such as ambulatory care. Therefore, self-collected finger-prick capillary blood sampling offers a suitable alternative to perform frequent measurements^3^. However, evaluations of such sampling remain limited. While microsampling techniques have demonstrated promising results for proteomics quantifications^4^, transcriptomics analyses on micro-samples are less common. It is thus important to determine whether venous and capillary samples yield comparable and reproducible results.

Several studies have compared changes in gene expression using capillary and venous blood samples with discordant reported findings^5,6^. However, more promising results have been presented elsewhere in terms of RNA quality^7^, agreement between finger-stick and venipuncture^8^ as well as clinical predictive capacity^9^. An ultralow-volume sampling procedure for self-collection and subsequent RNA preservation of a few drops of blood (50 µl) collected by a finger-stick has been successfully used to explore the response to COVID-19 vaccine^10,11^. In the context of COVID-19, a lot of data have been generated during hospitalization^12^ or in the post-hospitalization period^13,14^, but there are limited studies available examining the early stages of immune response in non-hospitalized patients^9,15-17^.

In the present work, we directly compared data derived from blood samples obtained simultaneously by venous puncture and ultralow-volume sampling collected by a fingerstick allowing the quantification of whole blood gene abundance applied to a cohort of COVID-19 patients followed-up at home in the framework of a clinical trial. We correlated changes in gene abundance to whole blood cell phenotype and serum biomarkers collected by venous puncture at fixed time points. Our results validate the finger-stick procedure, its feasibility at participant’s home, and provide insights about the dynamics of gene abundance changes during the early phase of mild COVID-19 infection.

## Methods

### Trial design and participants

The COVERAGE-Immuno study was embedded in the COVERAGE France trial, a phase-3, multicentre, open-label, randomized controlled trial of COVID-19 early treatment. The trial began in April 2020. The full original version of the protocol was previously published^18,19^.

COVID-19 positive outpatient participants who met all the inclusion criteria were randomly assigned at home or in an outpatient facility on day (D)0 to a control arm or one of the experimental arms ongoing at the time, and then visited on D1, D3, D5, D7, D9, D14, and D28.

Participants recruited at the sites were invited to take part in the COVERAGE-Immuno sub-study, including additional sampling for in-depth, repeated evaluation of the immunological markers and gene expression data (Supplementary Figure 1). Blood samples through veno-puncture were realized by a nurse at D0, D7 and D14 for the quantification of cell phenotype by flow cytometry on fresh blood (1 EDTA tube), serum analytes (1 dry tube) and gene expression (1 tempus tube). At D0, each patient was educated to self-sample by using the finger-prick at home (Supplementary Figure 1). Hence, self-collected finger-stick capillary blood samples were harvested at D0, D1, D3, D5, D7, D9, D12, and D14. Cotton swabs were stored in freezers and then collected weekly by a nurse.

### Ethics

The trial protocol was approved by a French Ethics Committee (CPPIDF1-2020-ND45). Written informed consent was obtained from all participants. The trial protocol was registered in the European database EudraCT (2020-001435-27) and on clinicaltrials.gov (NCT04356495).

### Quantification of serum analytes

Serum analytes were measured at baseline, 7 days and 14 days with samples collected during visits at home. Sixty-six Th1/Th2/activation/inflammation/apoptosis markers were measured in sera by Luminex assay, allowing the detection of 54 analytes with commercial kits (48-plex Bio-Plex Pro Human Cytokine screening Panel, Bio-Rad and 18-plex Human premixed Multi-Analyte magnetic Luminex Assay, R&D) according to the manufacturer’s instructions kits. Median fluorescence intensity for each sample was measured using the Bio-Plex 200 system (Bio-Rad, Marnes-la-Coquette, France). The Bio-Plex Manager software version 6.0, incorporating a weighted five-parameter logistic curve-fitting method, was used to calculate sample concentrations (in pg/ml). Twelve biomarkers showing more than 20% of undetectable data (below lower limit of quantification) were removed from the analysis. A total of 54 analytes was analysed.

### Cell phenotyping

All immunogenicity assays were performed on frozen peripheral blood mononuclear cells (PBMC). Immune phenotyping of the different cell populations was performed with an LSRII Fortessa 4-laser (488, 640, 561 and 405 nm) flow cytometer (BD Biosciences), and FlowJo software version 9.9.6 (Tree Star Inc.). i) CD4+ and CD8+T cells were analyzed for CD45RA and CC-chemokine receptor 7 (CCR7) expression, to identify the naive (CD45RA+CCR7+), central memory (CD45RA-CCR7+),effector memory (CD45RA-CCR7-) and terminal effector (CD45RA+CCR7-) cell subsets, and for co-expression of the activation markers HLA-DR and CD38; ii) B-cell subsets were analyzed with the CD19, CD21, CD27, CD38, immunoglobulin (Ig) D and IgM markers. Antibody-secreting cells are identified as CD19+ cells expressing CD38 and CD27; iii) NK cell subsets were analyzed using CD16, CD56 and CD57markers, as well as Ki 67 to define proliferating NK cells; iv) HLA-DR, CD33, CD45RA, CD123, CD141 and CD1c were be used to identify DC subsets, myeloid DC, plasmacytoid DC, DC1 and DC2.

### RNA sequencing

Gene expression profile was measured in Tempus samples at baseline, D7 and D14 as well as in cotton swab at the same time points, plus D3, D5, D9, D12.

Samples were analysed for gene expression profiles by mRNA sequencing. Total RNA was purified from whole blood using Tempus™ Spin RNA Isolation Kit (Invitrogen) and Globin mRNAs removed by GLOBINclear-Human Kit (Ambion). RNAs were then quantified with Quant-iT RiboGreen RNA Assay Kit (Thermo Fisher Scientific) before to be controlled for quality on the Bioanalyzer (Agilent). mRNA libraries were prepared by the TruSeq® Stranded mRNA kit according to Illumina protocol for Tempus whole blood recovery, and by the NEBNext Single Cell/Low Input RNA Library Prep Kit for Illumina for cotton prick samples. Libraries were sequenced on Illumina HiSeq 2500 V4 system. The sequencing depth were about 50 million reads and the fragment length of 1×100bp.

Quality control of the alignment was performed via MultiQC v1·13^42^. After this control stage, Trimmomatic v17·0·11^43^ was used to trim reads from sequences below a threshold, trim adapters and trim reads smaller than a defined length. The reads were aligned to the v44 human reference transcriptome (GRCh38.p14 from GenCode) using Salmon v1.8.0^44^ and quantified relative to annotation model “hsapiens_gene_ensembl” recovered from the R package biomaRt v2.58.0^45^. Finally, counts were normalized as counts per million.

### Statistical analysis

The concentration of 54 measured cytokines and the cellular responses assessed at D0, D7 and D14 were compared between baseline (D0) and the two other times separately using the Wilcoxon signed-rank test. We used the FDR method (Benjamini-Hochberg method) to adjust for test multiplicity.

For the transcriptomic part, differential expression analysis was carried out using the dearseq R package^27^ to analyze gene expression levels that differed between baseline (D0) and, respectively, D7 and D14 for finger-stick and tempus samples, separately. After the gene-level analysis, we focused on the gene set level with predefined gene sets: a fixed repertoire of 382 transcriptional modules that were thoroughly functionally annotated, “BloodGen3”^30^. From this, we conducted an analysis using the differential gene set function of the dearseq R package^46^ and visualized the results on fingerprint grids using the fold change (FC) (|FC| > 1·5). For finger-stick samples only, gene expression assessed at D0, D1, D3, D5, D7, D9, D12, D14 was analyzed for the gene and gene set levels with the same method previously described.

Cell deconvolution was inferred using CIBERSORT algorithm^37^ and the TIL10^38^ matrix reference. The estimated proportions were compared with the cell phenotyping using root-mean-square-error (RMSE), Pearson correlation and concordance correlation coefficient (CCC). These comparisons are shown in scatter plots and Blant-Alman plots.

Analyses were performed using R software (the R Foundation, Vienna, Austria; version 4·3).

## Results

### Participants and study feasibility

The COVERAGE France platform trial was designed to evaluate various treatment strategies in non-hospitalized patients with documented early COVID-19 and risk factors for aggravation of documented COVID-19. Patients with symptoms lasting for ≤7 days were recruited in the trial. The protocol has been already described^18,19^. An ancillary study was designed to explore host response in a subset of volunteering participants. After an initial training to the self-sampling method, the following sampling schedule was applied in volunteers : i) a venous puncture was performed by a nurse at participants’ home on D0, D7 and D14, and used for flow-cytometry cell phenotyping, quantification of serum analytes and RNA gene expression from whole blood; ii) a finger-prick self-sampling of capillary blood was performed at home by the participants themselves after education at D0, D1, D3, D5, D7, D9, D12, and D14, and used for the characterization of RNA gene expression (Supplementary Figure 1).

This ancillary study enrolled twenty-two participants from the COVERAGE trial recruited at Bordeaux site (mean age 65-years, 8 females) from August to November 2020. In median, patients were enrolled 4 (IQR 3-6) days after symptoms onset. They were randomized in a control arm (no treatment, n=10) or in intervention arms: (Imatinib; n=3; Telmisartan, n=9) (Supplementary Table 1). Two of them were subsequently hospitalized (at D5 and 10, respectively), none was admitted to intensive care unit, and none died during the 14 days of follow up post inclusion. A total of 156 finger-prick capillary samples have been performed and analyzed, 130/156 (83%) showing a good RNA integrity Number (RIN) > 6 (Supplementary Figure 2), demonstrating the feasibility of an intensive self-sampling at home despite acute sickness.

### Overview of early cell phenotype and serum biomarker profiles in patients with mild COVID-19

Cell phenotyping analysis showed an increase in the frequency of naïve CD4+ T cells at D7 which contrasts with the decrease of this cell population reported in more advanced patients^15,20^. At D14, we observed an increase of the expression of the activation marker HLA DR+ on CD4+ and CD8+ T cells as previously reported in patients with mild to severe disease by our group and others^21-24^. In addition, we observed an increase of the proportion of mDC without a decrease of pDC in contrast to more advanced patients^23^ (Figure 1A).

Measurement of serum markers revealed a low level of inflammation in these patients attested by the lack of elevation of markers such as IL-8 and TNF described in severe COVID-19 patients^25,26^. Similarly, IP10, IL-1b, PDL1, IL18 and FasLigand present at D0, decreased significantly at D14 exhibiting a profile of transient burst as previously described in mild infection^9^ and which differs from severe infections. Of note, IFN levels remained stable from baseline (Figure 1B). Therefore, patients enrolled in the COVERAGE ancillary study exhibited clinically and biological profiles consistent with a mild COVID-19^15^ (Supplementary Figure 3). In this context, we sought to extend here investigations on the dynamics of early molecular immune response to SARS-CoV-2 by conducting a thorough analysis of additional markers, measured more frequently, leveraging the gene expression abundance data obtained from finger-stick capillary samples.

**Figure 1.**
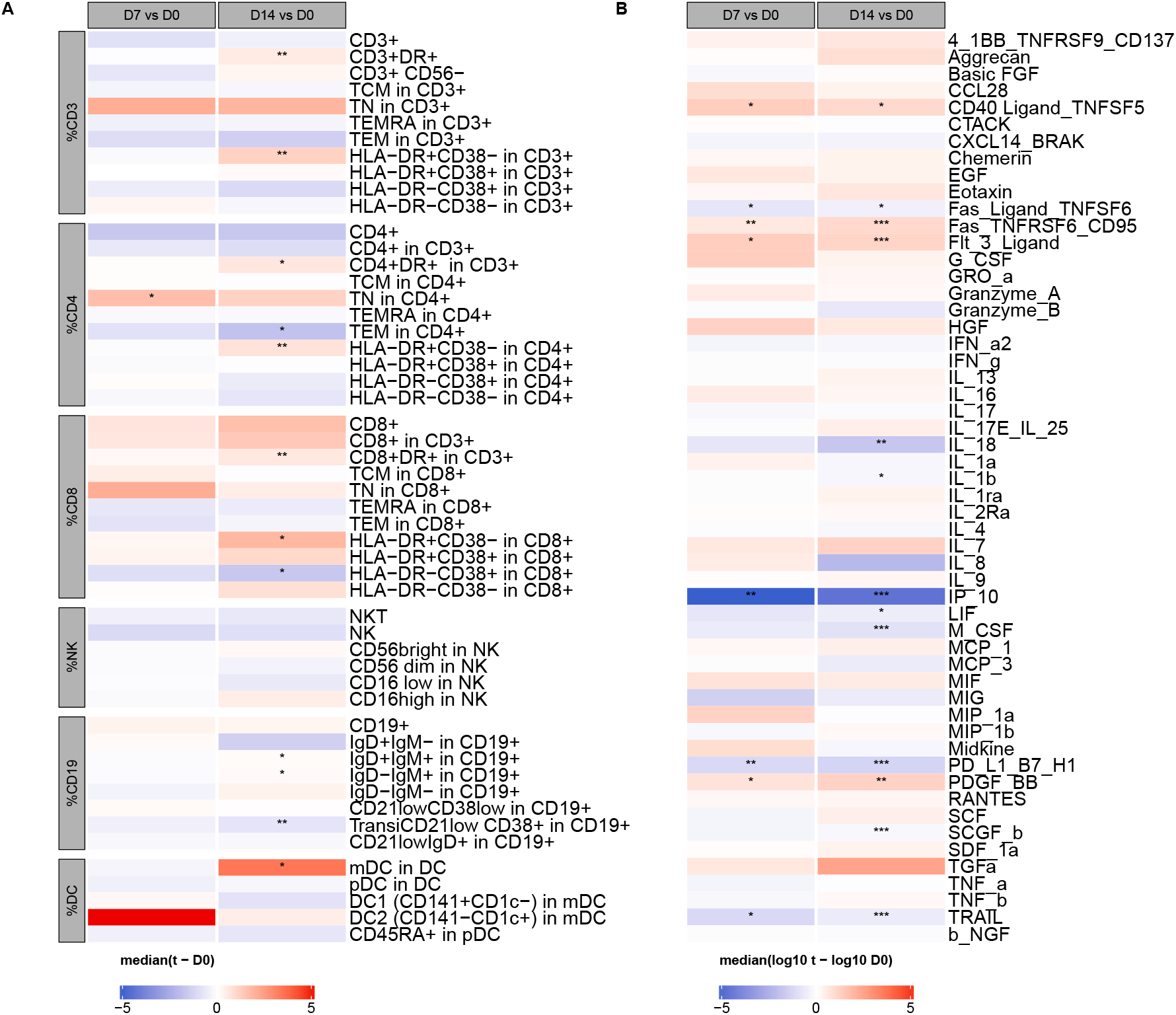
Cellular and serum cytokine profiles over time. **A**. Heatmap showing median difference of proportion of cells between day 7 (D7), day 14 (D14) and baseline. **B**. Heatmap showing median log10 difference between day 7 (D7), day 14 (D14) measures and baseline for serum cytokines. Wilcoxon test after FDR adjusted p-value: *<0·05, **<0·01, ***<0·001.

### Comparison between finger-stick and venous sampling demonstrated a good concordance and pathways enrichment

First, we compared the abundance of gene expression at D7 and D14 to D0 in samples collected by finger-stick or venous blood puncture. The intraclass correlation of gene abundance between the two sampling methods was excellent (r = 0·74, Figure 2A). In the Bland and Altman plot (average of the two measures on x-axis and difference between the two measures on the y-axis), 93% of the measures were included in the mean range ± 1,96 standard deviations demonstrating an excellent agreement between the two measurements of gene abundance (Figure 2B).

**Figure 2.**
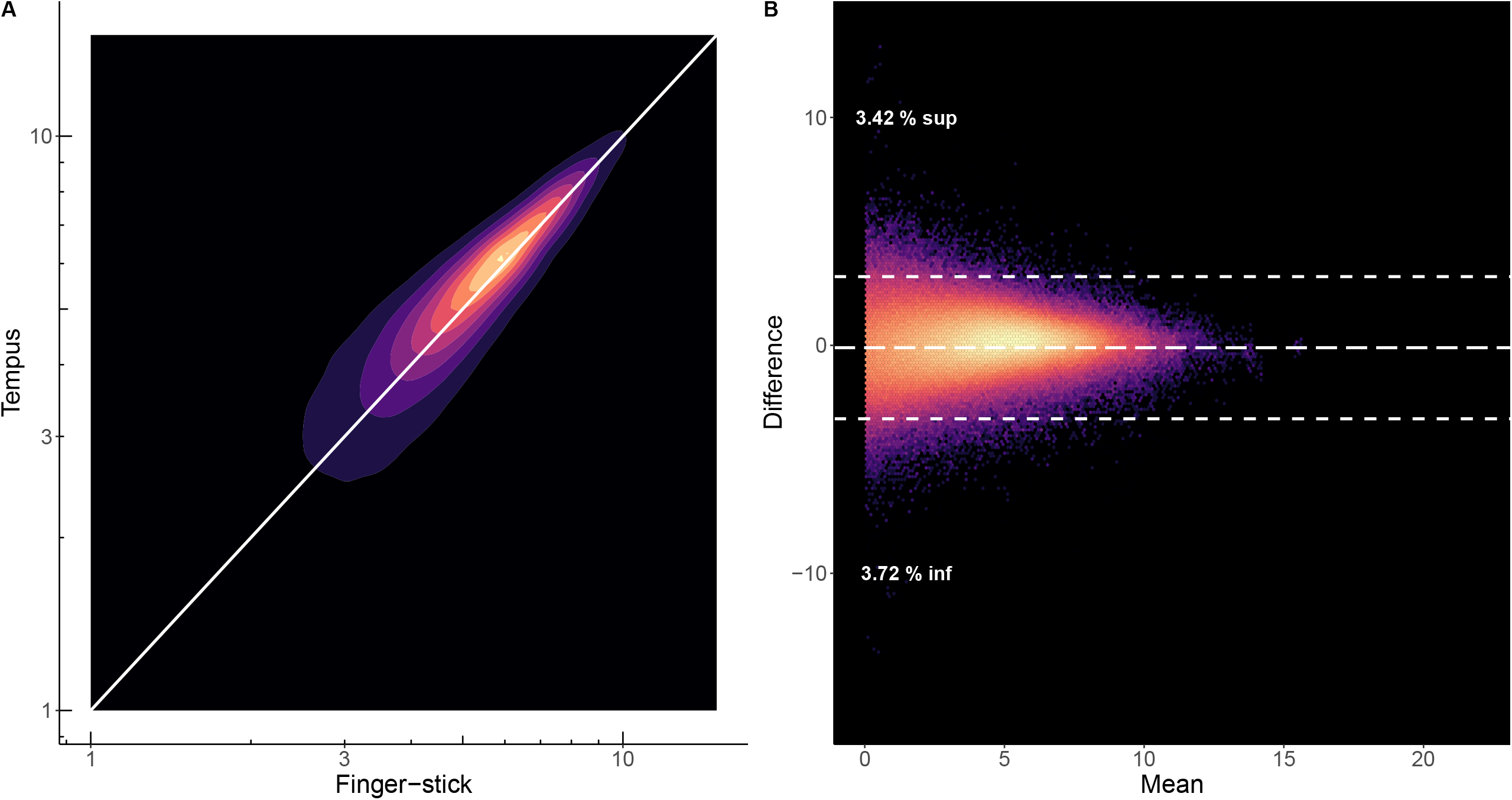
Agreement between the RNA sequencing normalized data of COVID19+ outpatient measured by finger-stick and Tempus sampling (all genes, all time points) **A**. Correlation between RNA sequencing measured by finger-stick and Tempus (r = 0·74). **B**. Bland and Altman plot for agreement between RNA sequencing measured by finger-stick and Tempus (average of the two measures on x-axis and difference between the two measures of the y-axis). 93% of the measures are included in the range.

Then, we looked at the differential abundance of the genes between D0 and D7 and D14 provided by the two technics using the dearseq methods^27^(Supplementary Figure 4). At D7, 265 and 223 genes were differentially abundant in finger-stick and venous blood samples, respectively. The differentially abundant genes increased at D14, to reach 352 and 690 genes abundant in finger-stick and venous blood samples, respectively. Among the top 20 genes differentially expressed at D7, 10 of them were similar regardless the sampling method: *HESX1, OTOF, LAMP3, PI3, CCL8, CCL2, TMEM255A, SIGLEC1, IFI44L, RSAD2* (Supplementary Figure 4). Interestingly, among these top 20 genes, RSAD2 or viperin and IFI44L are interferon-stimulated genes, with strong anti-viral activity. Besides, CCL8 (MCP-2) and CCL2 (MCP-1) are monocyte chemoattractant cytokines that are shown significantly stimulated during COVID-19 infection. At D14, although the signal seems lower in the finger-stick samples, 11 over the top 20 genes were common to the two sampling methods (*KCTD14, HESX1, OTOF, IFI27, SIGLEC1, LAMP3, IFI44L, RSAD2, ZNF496-DT, LY6E-DT, SPP1*).

These data provide a possible answer to previous studies raising concerns about the interest of finger-stick gene expression analysis because of the absence of agreement with venous blood sampling^6^. Although biological explanations could be hypothesized according to the previously published differences in cell frequencies^28^ or cytokines^29^, a technical bias should be discarded before implementing finger-stick analysis. For example, we re-analyzed previously published microarray data^5^ for which Stein et al^6^ originally claimed there was a strong divergence between the two sampling techniques. We show instead that this discrepancy could rather be linked to a batch effect, that is absent in our present study. Therefore, our results reinforce the interest and reliability of finger stick method, and also underscores the requirement of checking potential bias in the generation of sequences.

To confirm the interest of the finger-stick approach, we extended the comparison of the results between the two sampling methods in term of geneset enrichment (Figure 3) which are group of genes defined *a priori* using other datasets^30^. We hypothesized that this comparison would be more relevant as it is less sensible to the variation of the signal at the gene level. Analyses were carried out using a fixed repertoire of 382 transcriptional modules (BloodGen3) and characterized functionally^30^. The gene composition of the modules and the functional annotations are provided here https://drinchai.shinyapps.io/BloodGen3Module/. “Module aggregate” are sets of modules that showed consistent abundance profiles across reference set of 16 disease cohorts that were used for the construction of the BloodGen3 repertoire^30^. Although there were more modules that appeared to have an evolution of their abundance in the venous blood sampling method as compared to the finger stick approach, the agreement between the methods for the modules with a clear homologous change of the gene abundance was perfect: the abundance of genes from module M10·4 (*BPI, CEACAM6, CEACAM8, CTSG, DEFA1, DEFA3, DEFA4, ELA2, LOC653600, LOC728358, LTF, MPO, OLFM4*) annotated “Neutrophils activation” was found significantly increased at day 7 and day 14 with the two sampling methods. The abundance of the five on six modules belonging to the module aggregate A28 annotated “Interferon” was found consistently decreased with the two sampling methods at the two time points as well as the module “Type 1 Interferon” including *IFI, IFIT, OAS, EPSTI1, HERC5, HES4, IFI44, IFI44L, IFI6, IFIT1, IFIT3, ISG15, LY6E, MX1, OAS1, OAS2, OAS3, OASL, RSAD2, XAF1*) genes (Supplementary Figure 5-6). The aggregate annotated “Erythroid cells” presented an increased abundance at D14 only but consistently with the two sampling methods. Consistently, the two sampling methods, albeit with a weaker signal, showed also the modules “cell cycle”, “inflammation”, “protein synthesis” and “monocytes” as differentially abundant at D7 and D14 as compared to D0.

**Figure 3.**
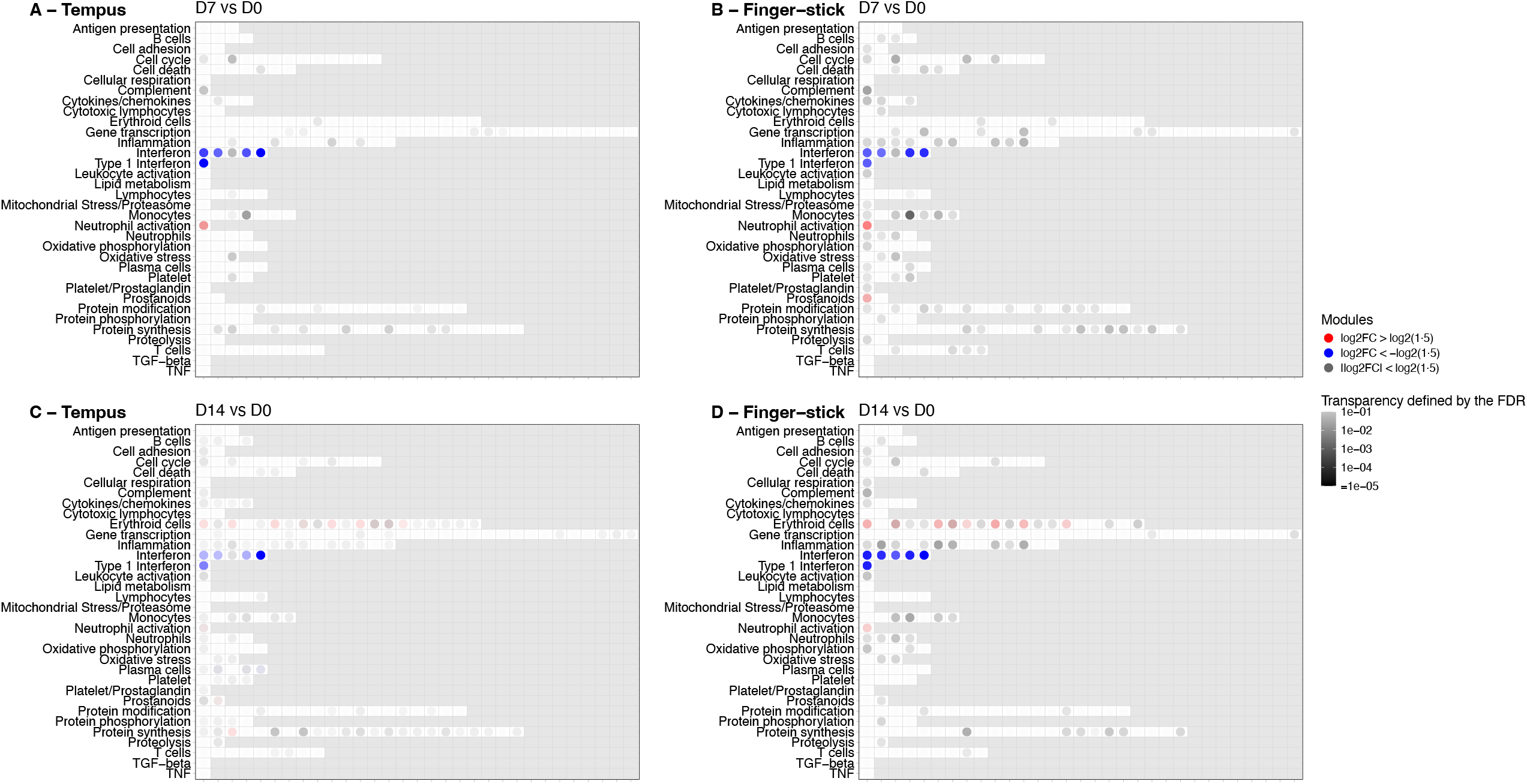
Description of the enriched gene sets at day 7 post inclusion measured in venous blood and finger-stick samples at day 7 (A, B) and day 14 (C, D), respectively, in the 22 COVID19+ outpatient of the COVERAGE immune study. Modules that are colored presented a differential abundance compared to baseline with absolute FC > 1·5 and FDR < 0.01.

These results showed a high consistency in the enrichment of gene sets between the two sampling methods and confirmed signals such as neutrophils activation, interferon signaling, erythroid cells and inflammation pathway at early stages of mild COVID-19. Several of these changes are in agreement with those previously described and associated with severe disease by our group, and others^31^. However, these changes were described at later time points after disease onset (one month) and persisted several months in the settings of long COVID^31^. Therefore, while the persistence of these abnormalities is well described in hospitalized patients with severe disease, the dynamics of these immune disturbances during the early stages of the infection in outpatients is not fully described.

### The high temporal resolution transcriptomics analysis using finger-stick sampling revealed a more precise dynamics of gene abundance from key pathways

As described above the dynamics of changes in gene abundance was at first limited to the resolution of only two time points (D7 and D14) due to the constraint of venous blood sampling. The finger-stick sampling allowed us to better decipher early gene changes every two days from D0 to D14. We showed that patients with mild COVID-19 experienced an interferon response with an increased in related gene abundance since D1 with a rapid decrease from D3 to D9 (Figure 4A-B, supplementary Figure 7 A-B). In contrast, neutrophil activation attested by increased abundance of genes from the neutrophil module increased from D1 and persisted until D12 (Figure 4 C-D, supplementary Figure 7 C-D). In regards of the aggregate A37 (“Erythroid cells”), the dynamics of the gene abundance was very different with an increase starting on D3 to D14 (Figure 4 E-F, supplementary Figure 7 E-F).

**Figure 4.**
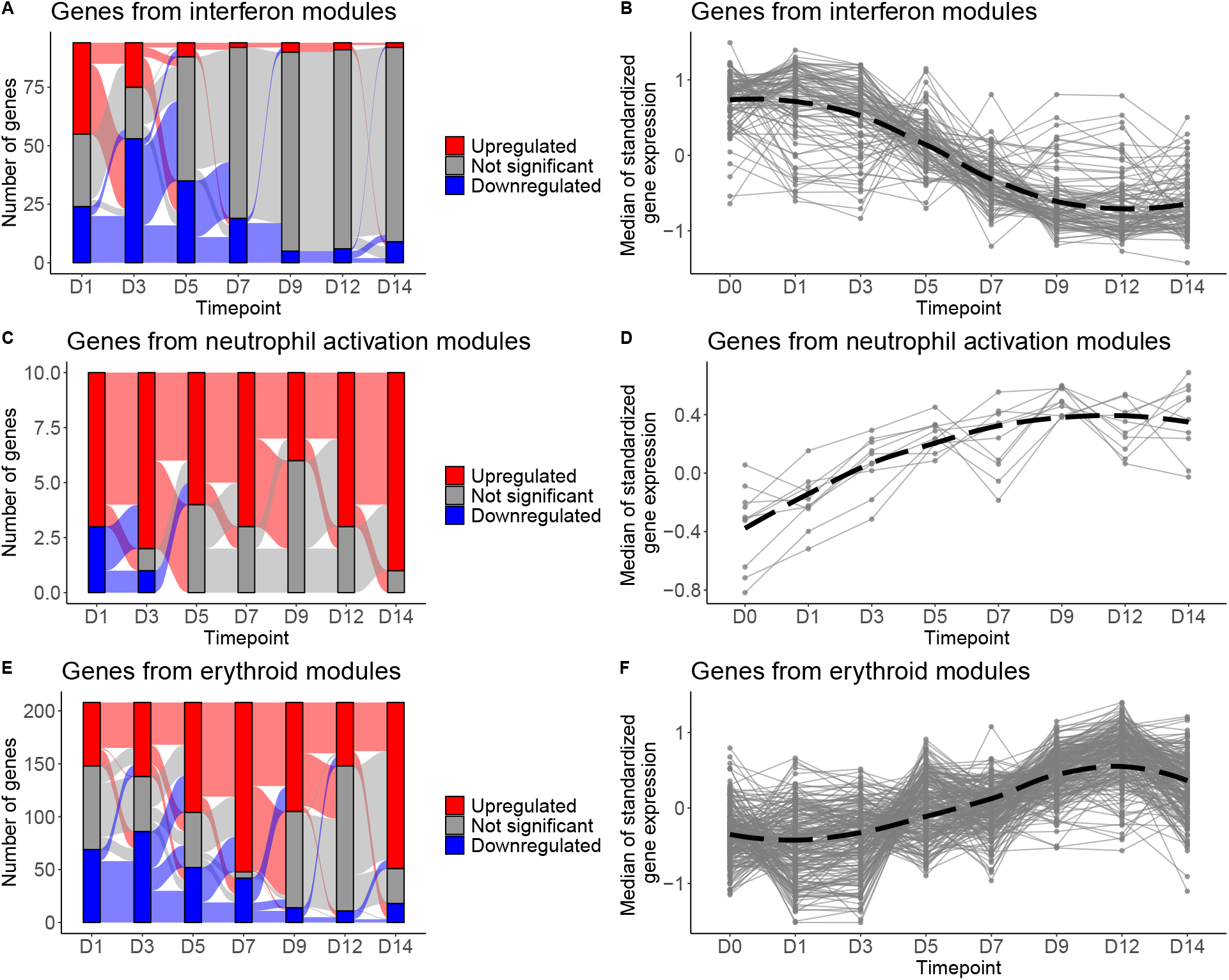
Change of gene abundance every two days associated to interferon regulation, neutrophil activation and erythroid cells using finger-stick capillary sampling. Genes differentially expressed and evolution of abundance of genes of interferon (**A-B**), neutrophil (**C-D**) and erythroid (**E-F**) modules over time (each time point compared to day 0).

Overall, we show here that repeated finger-stick sampling revealed a more complex and non-linear dynamics in changes in gene abundance of critical pathways at the early stages of mild COVID-19. For instance, we showed an early interferon response may have been beneficial, which is consistent with the outcome of these patients^32^. This response waned in few days (beyond day 3) indicating that studies looking at later time points may miss these early changes in gene expression^33,34^ with the exception of human challenge studies that reported the same results^17^. Globally, repeated at-home measurements allowed to link a favorable clinical evolution of COVID-19 in elderly patients to a quick resolution in a couple of days of the immune response to the infection.

### Correlation between whole blood gene expression and variations in cell populations and cytokine levels

Several deconvolution algorithms have been proposed to infer blood transcriptomic changes to variations in various type of cells^22,35,36^. Here, we used CIBERSORTx^37^ algorithm with TIL10^38^ reference matrix. We found a good correlation between gene abundance, regardless the sampling method, and the frequency of B and CD8+ T lymphocytes as assessed by flow cytometry (Figure 5A-B). However, none of the sampling methods provided a good prediction between gene abundance and the frequency of CD4+ T cells, NK cells or neutrophils (Figure 5C-E). More interestingly, we looked at the concordance between gene expression and two markers of inflammation; i.e., IL-1b and IP-10 (Figure 6A). We showed an agreement with a high precision in the temporal trends of these markers when measured by Luminex. Moreover, the two-days finger-stick sampling allowed to finely analyze these dynamics showing an increased and stable expression over the first 3 days before dropping in a nearly linear shape over time (Figure 6B). The rapid observed decrease of IL1b is in agreement with the decrease of circulating inflammatory monocytes as reported in human challenge studies^17^.

**Figure 5.**
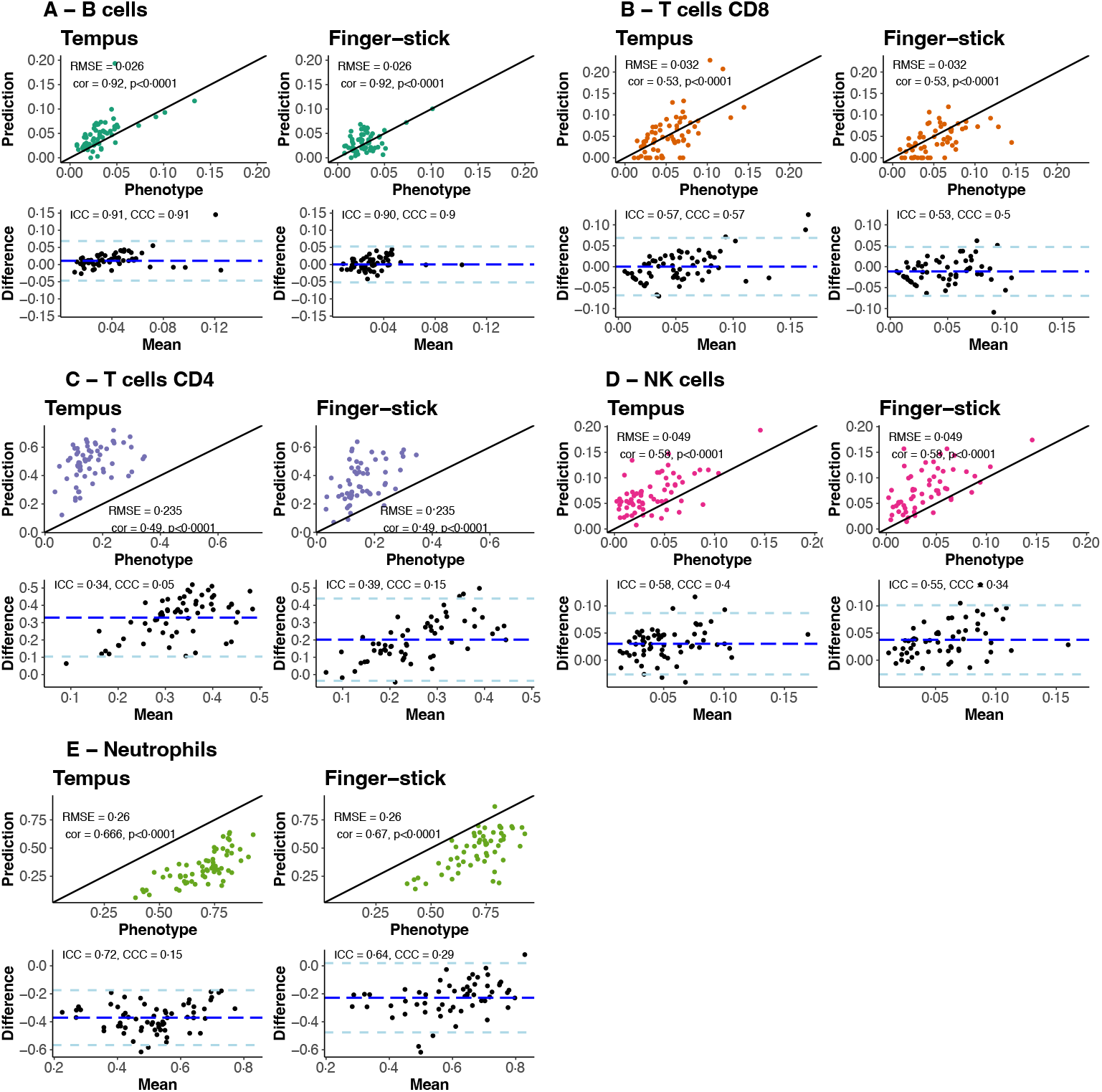
Correlation between gene abundance measured by Tempus (left) or finger-stick (right) and cell abundance (Flow cytometry) for B cell, T cell CD8, T cell CD4, NK cells and Neutrophils respectively.

**Figure 6.**
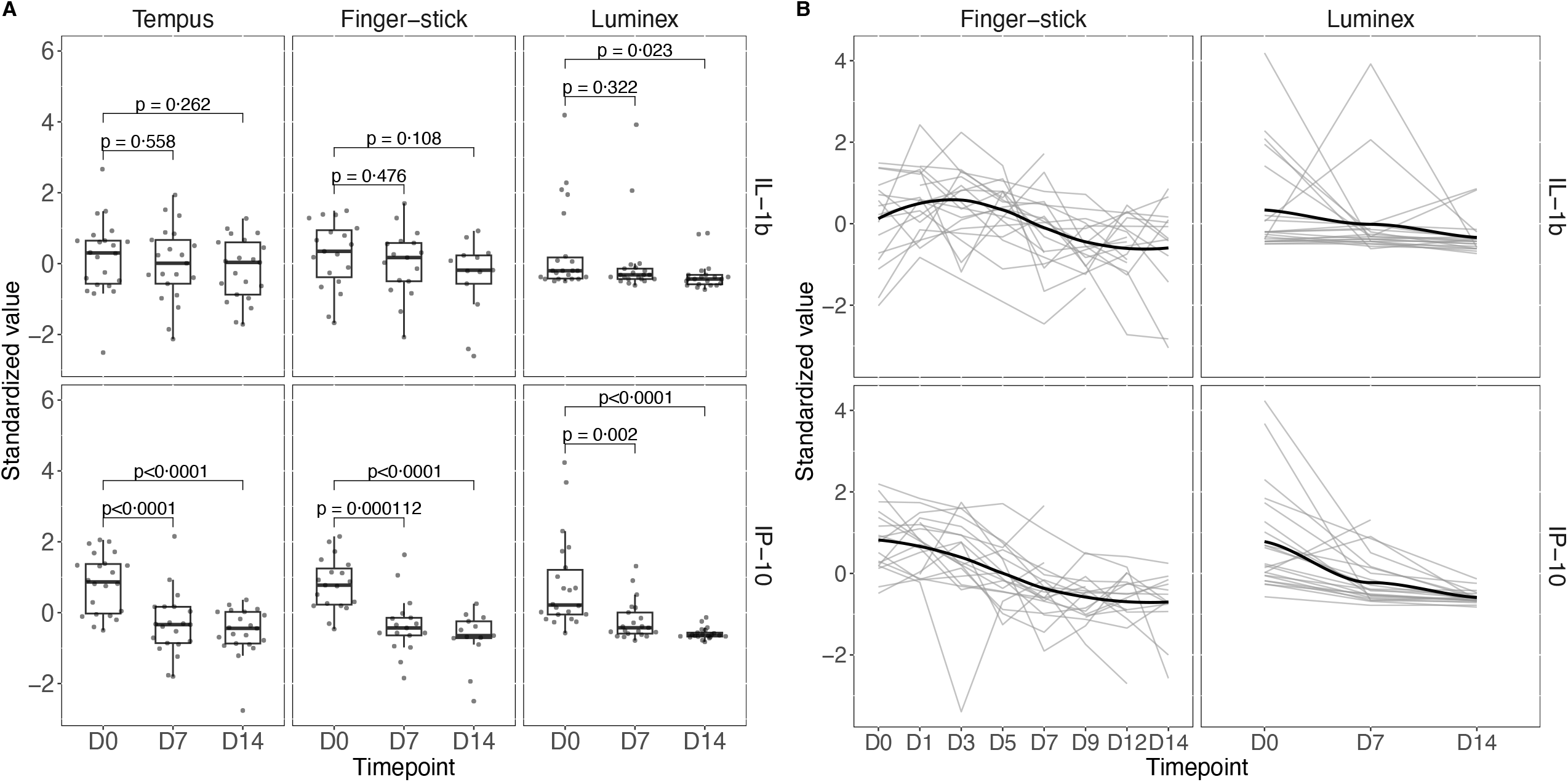
Longitudinal analysis of inflammatory markers (IL-1b, IP-10) measured by RNA sequencing (tempus and finger-stick) and Luminex (serum) respectively.

## Discussion

Overall, the comprehensive data generated in this study provide a novel approach to interrogate the molecular mechanisms underlying host responses at early stages of COVID-19 in a population of patients at high risks of aggravation defined by age and comorbidities. We showed that obtaining reliable and reproducible gene expression data from a fingerstick has obvious advantages as compared to specific time points guided by the constraint of venous punctures. These new data exploring early responses to SARS-CoV-2 infection extend previous reports which successfully used finger-stick sampling to characterize auto-immunity^39^, as well as the response to infection^40^ or to vaccination^41^. We showed here that this sampling approach could be useful to identify pathways and to characterize dynamics of gene expression that are missed by more targeted studies with predefined later time points. One interesting information provided by this study, thanks to repeated measurements, is that earliest stages of the SARS-CoV-2 mild infection are characterized by the same pathways described in patients with severe diseases but with a quick resolution. In addition, we showed a correlation between gene abundance and deconvolution of various cell frequencies as well as serum biomarker dynamics which represent an opportunity for clinical research and to better decipher SARS-CoV-2 physiopathology.

Our study has limitations. One of those is the low number of participants. Despite this, we were able to demonstrate a statistically high concordance between the two sampling methods in the analysis in gene abundance. The objective of our study was to demonstrate the feasibility of at-home sampling in the context of acute SARS-CoV-2 infection, we were not able to analyze the impact of therapeutic interventions (Imatinib, Telmisartan). We showed some early features of SARS-CoV-2 infection in elderly patients infected in 2020-2021 when the COVERAGE trial was launched. Whether these earliest molecular and immunological profiles are still relevant with subsequent variants remain to be investigated.

The use of finger-prick tests in clinical research represents a genuine opportunity to transform this field by identifying early biomarkers following a diagnosis or intervention, conducting intensive studies of their dynamics, and doing so with samples collected by the participants themselves. Precision medicine could see a new dawn with this technology.

## Contributors

RT, EL, BH, YL designed the study. XA, DM enrolled and followed the patients and recorded clinical data. HH, MS, IP, ABM have performed the biological analyses. MH, BH, MP, EL, RT performed the statistical analysis. RT, YL and EL drafted the first version of the manuscript. All authors revised the manuscript critically for important intellectual content and approved the final version before submission. EL and HH contributed equally to this work.

## Acknowledgments

This work was supported by INSERM and the Investissements d’Avenir program, the Vaccine Research Institute (VRI), managed by the ANR under reference ANR-10-LABX-77–01, and the CARE project, funded from the Innovative Medicines Initiative 2 Joint Undertaking (JU) under grant agreement No 101005077. The JU receives support from the European Union’s Horizon 2020 research and innovation program, the EFPIA, and the Bill & Melinda Gates Foundation, Global Health Drug Discovery Institute, University of Dundee.

We thank all the persons who participated in the trial, and the following members of the COVERAGE France Study Group: Duvignaud A, Bez C, Bironneau E, Collomb E, Contamin A, Dubourdieu C, Faure N, Galinski M, Gibaud P, Gil-Jardine C, Guillot-Warin J, Lebouc C, Leger A, Lengline V, Loizeau C, Mayenc M, Merle N, Nadiri-Kahraman H, Nguyen D, Odorico M, Onaisi R, Sacher F, Scandella J, Velardo F, Wiet A.

## Sharing statement

### Data availability

Data supporting this study can be accessed on BioStudies database - ebi.ac.uk (E-MTAB-15066).

### Code availability

Code related to this publication is available at https://doi.org/10.5281/zenodo.14447221

