## Supplementary Materials for "Self-Collected Finger-Prick Blood for Gene Expression Profiling: Unveiling Early Immune Responses in Mild COVID-19"

### Supplementary information

COVERAGE France study group, Supplementary Figures. 1–7, Table 1, methods, results, Supplementary Note 1

**COVERAGE Study group**

- Sponsor (CHU Bordeaux):

G. Duluc (head), D. Arma, P. Beaufrère, J. Belcastro, T. Brice, P. Cassai, S. Desjardins, J. Durrieu, S. Georgevail, A. Gimbert, S. Marchi, V. Marty, F. Nacka, R. Pinilla, P. Poulizac, S. Regueme, M. Rousset, C. Roussillon, F. Salvo, J. Soria, S. Vautrat

- National Trial Coordination Unit:

***CIC-EC 1401/EUCLID***: L. Richert (head), F. Allais, S. Canete, N. Chaghil, S. Daoui, G. Dupouy, L. Esterle, V. Favreau, A. Gelley, S. Gillet, L. Hardel, M. Kanté, E. Lhomme, S. Martiren, L. Moinot, L. Pinoges, E. Rouch, C. Schwimmer, R. Sitta, M. Termote, C. Wallet, L. Wittkop

***MEREVA:*** X. Anglaret (head), E. Balestre, A. Beuscart, C. Bonnier, C. Cazes, C. Chazallon, G. Clouet, M. Daures, D. Gabillard, G. Habiyambere, V. Journot, M. Loniewski, S. Karcher, J. Le Carrou,O. Marcy, V. Murat, J. Orne Gliemann, M. Plazy

- National Coordinating Pharmacy:

S. Djabarouti (head), B. Ghezzoul, L. Delignac, P. Etienne, V. Fulda, M. Gigan, C. Langlade, J. Le Chanjour, P. Marque, P. Mora, C. Plessis, B. Sourisseau

- National Coordinating Laboratories:

**Bordeaux:** I. Pellegrin (head), S. Bouchet, C. Cognet, I. Garrigue, J. Jeanpetit, M-E. Lafon, A. Pouzet, A. Tarricone, P. Trimoulet, A. Voldoire

**Créteil:** C. Lacabaratz (head), H. Hocini

- Bordeaux Study center:

***Investigating physicians*:** X. Anglaret, A. Duvignaud (Principal Investigator), C. Bez, E. Bironneau, E. Collomb, A. Contamin, C. Dubourdieu,, N. Faure, M. Galinski, P. Gibaud, C. Gil-Jardine, J. Guillot-Warin, C. Lebouc, A. Leger,V. Lengline, C. Loizeau, M. Mayenc, N. Merle, H. Nadiri-Kahraman, D. Nguyen, M. Odorico, R. Onaisi,T. Pistone, F. Sacher, J. Scandella, F. Velardo, AS. Wiet

***Coordination*:** X. Anglaret (head), T. Boudon, O. Bouissière, R. Brégéras, N. Broennec, S. Condé, C. Gazille, C. Grenier, G Kouame, C. Martins-Calado, J-B. N’Takpe, Z. Pascual

- Dijon Study center:

***Investigating physicians*:** L. Piroth (Principal Investigator), S. Court Devilliers, J. Darley, F. Ducherpozat, N. Eberard, JL. Faure, H. Portier, R. Rochelet, C. Ruffino, R. Thevenoud

***Coordination*:** C. Binquet, TT. Creusvaux-Nguyen, N. Desbiolles, A. Grattard, A. Lamotte Felin, L. Rossye, C. Schaeffer, F. Silvestre, C. Simonel,

- Nancy Study center:

***Investigating physicians*:** B. Lefèvre (Principal Investigator), G. Baronnet, E. Baux, C. Di Santolo, M. Ferry, F. Goehringer

***Coordination*:** C. Daguin, E. Dauchy, M. Gilg, P. Rossignol

- Toulouse Study sites:

***Investigating physicians*:** J. Dupouy (Principal Investigator site 1), A. Boucault (Principal Investigator site 2), L. Gimenez (Principal Investigator site 3), C. Burguier, M. Couderc, C. Fradet, A. Gervais, J. Lavergne, C. Landon, S. Mathe, B. Ortala

***Coordination*:** N. Ainaoui, S. Auriac, S.Bras , S. Flasquin, C. Fraysse, M. Gabriel, P. Gauteul, J. Germain, F. Gross, C. Lebely, C. Riviere, D. Robert, C. Thalamas

- Bastia Study center:

***Investigating physicians*:** T. Darnaud (Principal Investigator), D. Poitrenaud

***Coordination*:** I. Giusti, S. Provent

- Montpellier Study center:

***Investigating physicians*:** A. Makinson (Principal Investigator), A. Montoya-Ferrer

***Coordination:*** J. Battery, N. Coux, L. Crantelle, F. Galtier, C. Martin, MC Picot

- Paris Study center 1 : Collège National des Généralistes Enseignants (CNGE)

***Investigating physicians*:** J. Le Bel (Principal Investigator), J. Chastang, A. Nguyen, O. Saint-Lary

***Coordination*:** I. Giraud

- Paris Study center 2 : Groupe Hospitalier Paris Saint Joseph (GHPSJ)

***Investigating physicians*:** JM. Naccache (Principal Investigator site 1), J. Jouveshomme, E. Devaud (principal investigator site 2)

***Coordination*:** N. Ben Nasr, R. Monkam, E. Sacco, S. Rulle

- Paris Study center 3 : Institut de Médecine et d’Epidémiologie Appliquée (IMEA)

***Coordination*:** R. Landman (principal investigator), K. Amat, A. Benalicherif, B. Sylla

- Nantes/Angers Study center:

***Investigating physicians*:** C. Begue (Principal Investigator site 1), C. Rat (Principal Investigator site 2),

C. Bouve, C. Huard, S. Morgand

***Coordination*:** E. Guegan, D. Fairier, A. Loiez

- Others contributors:

T. Alcouffe, V. Briand, S. Coudray, D. Charles, H. Jacquet, C. Levy- Marchal, J. Raude, P. Vellozzo, P. Whithenay

- Scientific Advisory Board:

**Voting members*:*** X. de Lamballerie (Chair), X. Anglaret, L. Atlani-Duault, C. Begue, F. Chauvin*, J. Chastang, D. Darmon, T. Darnaud, P-L. Druais, V. Dubee, J. Dupouy, A. Duvignaud, A. Gimbert, V. Journot, R. Landman, D. Lebeaux, B. Lefèvre, E. Lhomme, A. Makinson, D. Malvy, F. Mentré, J-F. Michel*, M. Molimard, JL. Montastruc, JM Naccache, J. Orne Gliemann, L. Piroth, C. Rat, L. Richert, C. Roussillon, O. Saint Lary, R. Thiebaut, L. Weiss, L. Wittkop

**Non-voting members*:*** S. Bouchet, C. Cazenave, S. Conde, A. Cremer, S. Djabarouti, C. Gil Jardine, L. Hardel, J-P. Joseph, M-E. Lafon, J. Le Bel, L. Letinier, S. Marchi, L. Moinot, A. Montoya-Ferrer, JD. Nguyen, R. Onaisi, I. Pellegrin, T. Pistone, D. Poitrenaud, T. Schaeverbeke

- Data Safety Monitoring Board:

D. Costagliola (Chair), E. Bellissant, G. Gavazzi, C. Locher, A-M. Taburet, P. Tattevin, S. Walker

**Supplementary Methods**

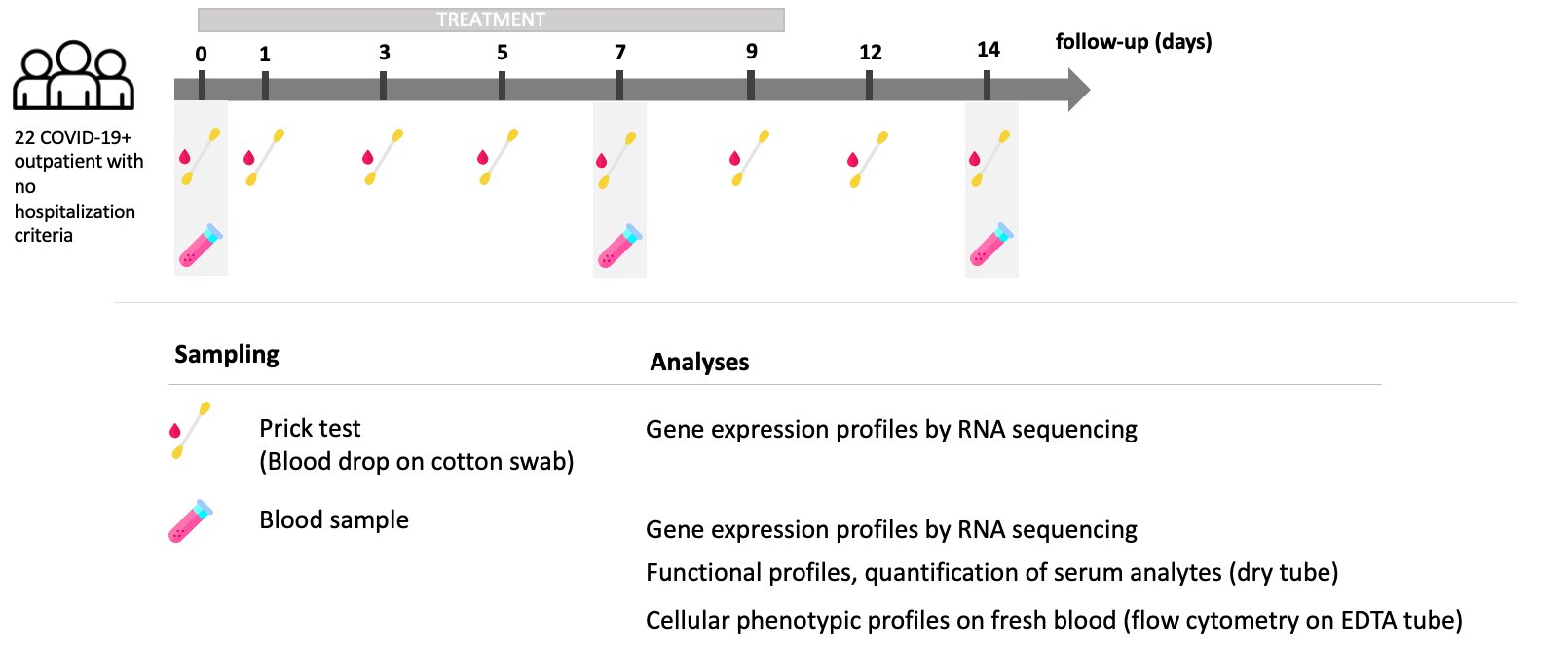

**Supplementary Figure 1. Design of COVERAGE immunological sub-study**

### Supplementary results

**Supplementary Table 1. Description of the 22 COVID-19+ outpatients included in the COVERAGE immunological study.** A total of 22 participants in Bordeaux site have been included in COVERAGE immuno. The median age was 65 years (inter-quartile range (IQR): 63-73) and 14 (64%) were male. The median delay of symptoms onset at the inclusion was 4 days (IQR 3-6). On the 22 participants included, 10 (45%) were randomized in the control group, 9 (41%) in the Telmisartan arm and 3 (14%) in the Imatinib arm, which were the two experimental arms opened during. At day 14, only 2 (9%) participants were hospitalized, none in intensive care unit, and no participant died.

| **Variable** |  | **All (N=22)** |
| --- | --- | --- |
| Age (years) - Median(IQR) |  | 65 (63 – 73) |
| Male - Nbr/Total nbr (%) |  | 14/22 (64) |
| Hospitalization - Nbr/Total nbr (%) |  | 2/22 (9) |
| Death - Nbr/Total nbr (%) |  | 0/22 (0) |
| Delay symptoms onset - Median(IQR) |  | 4 (3 - 6) |
| Arm Azinc Vitalité - Nbr/Total nbr (%) |  | 10/22 (45) |
| Arm Imatinib - Nbr/Total nbr (%) |  | 3/22 (14) |
| Arm Telmisartan - Nbr/Total nbr (%) |  | 9/22 (41) |

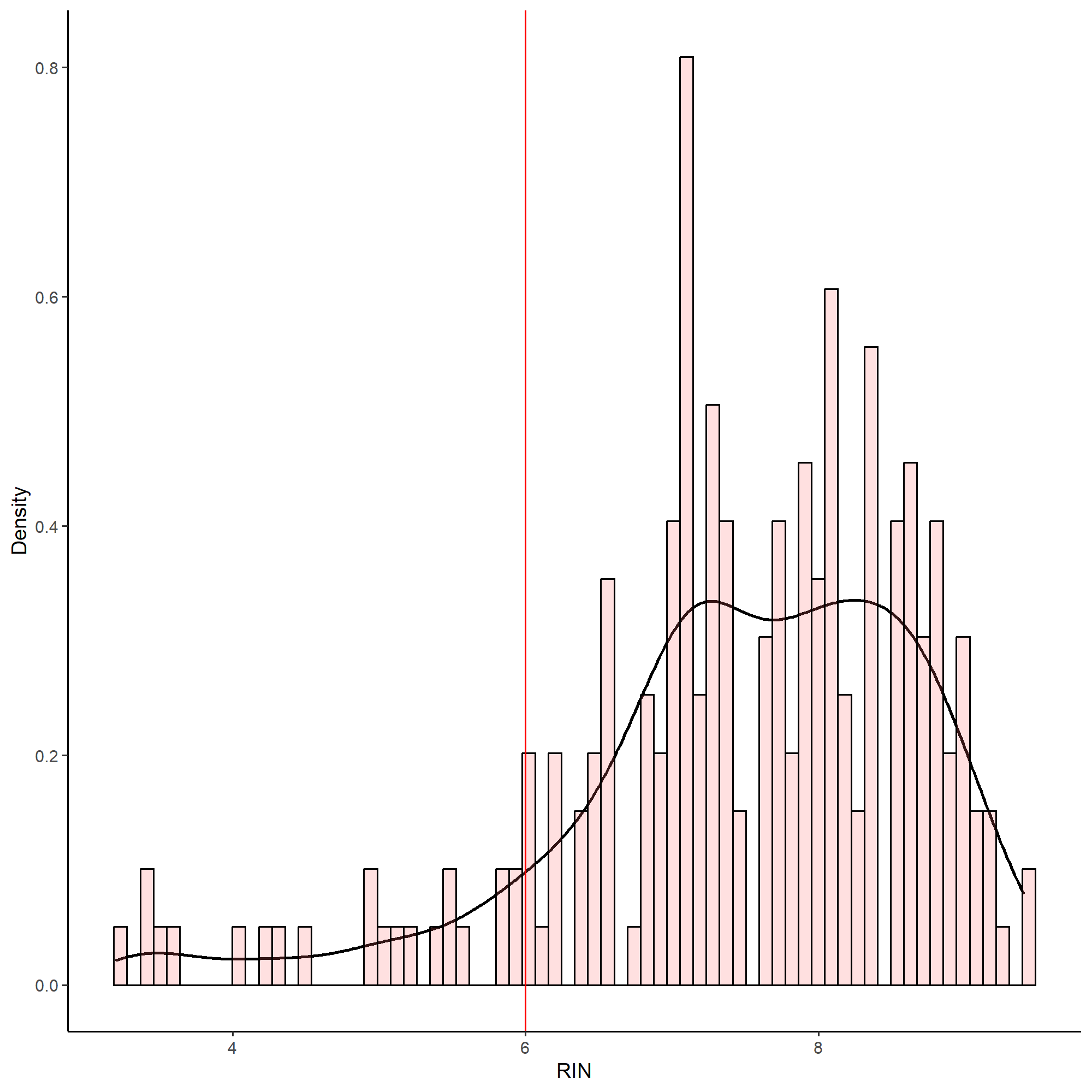

**Supplementary Figure 2. RNA integrity number (RIN) distribution for prick test samples.**

**A**

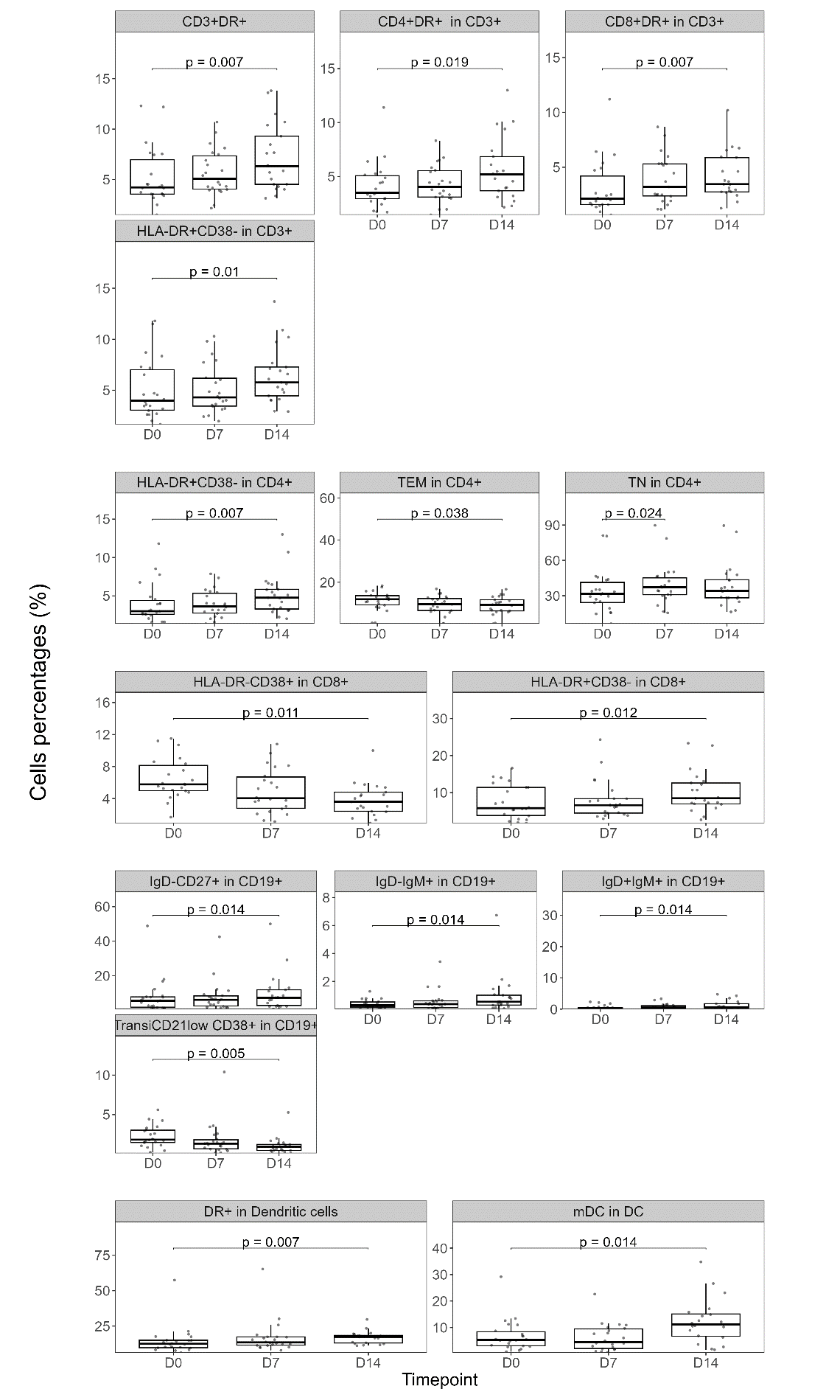

**B**

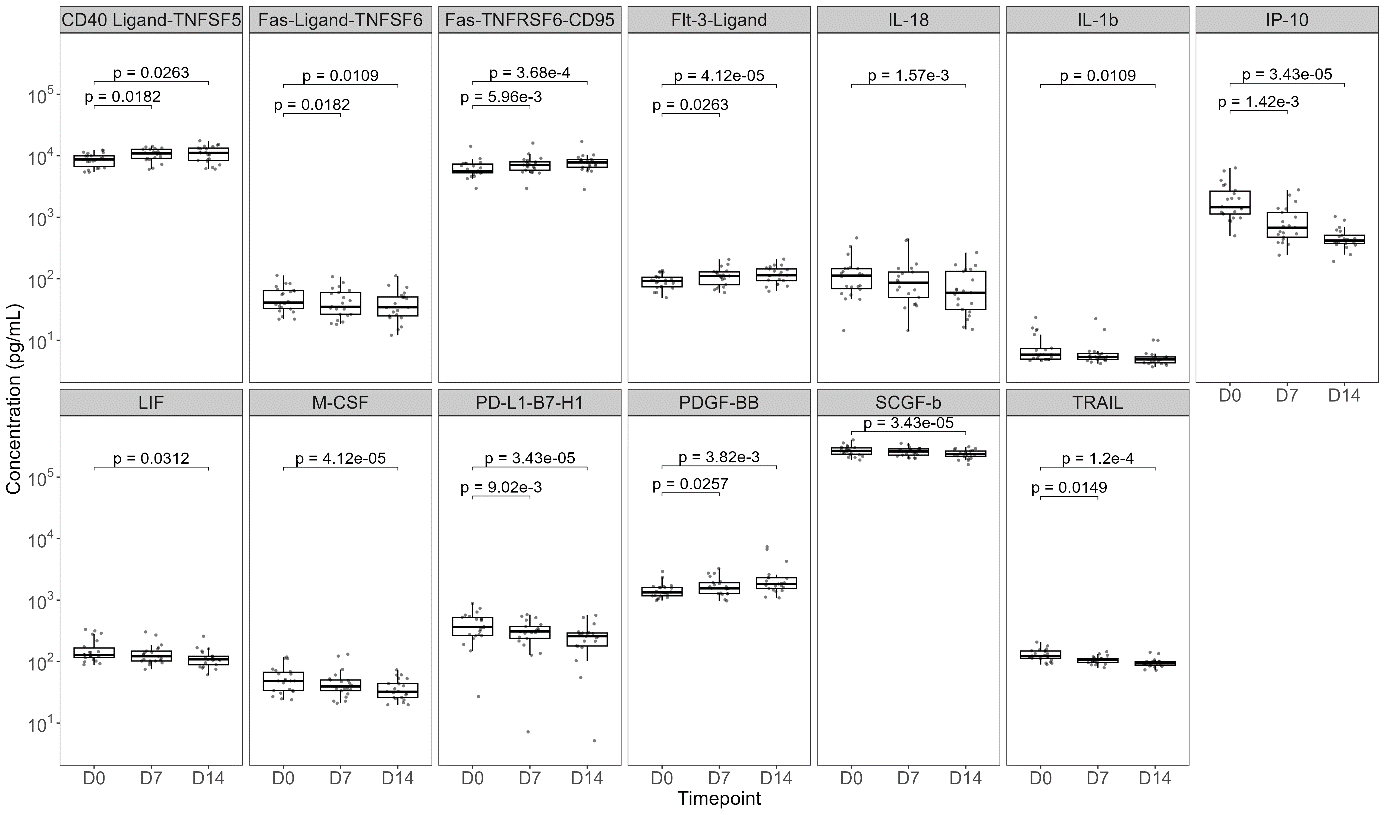

**Supplementary Figure 3.** **Longitudinal analysis measured by Flow cytometry (A) and Luminex (B).**

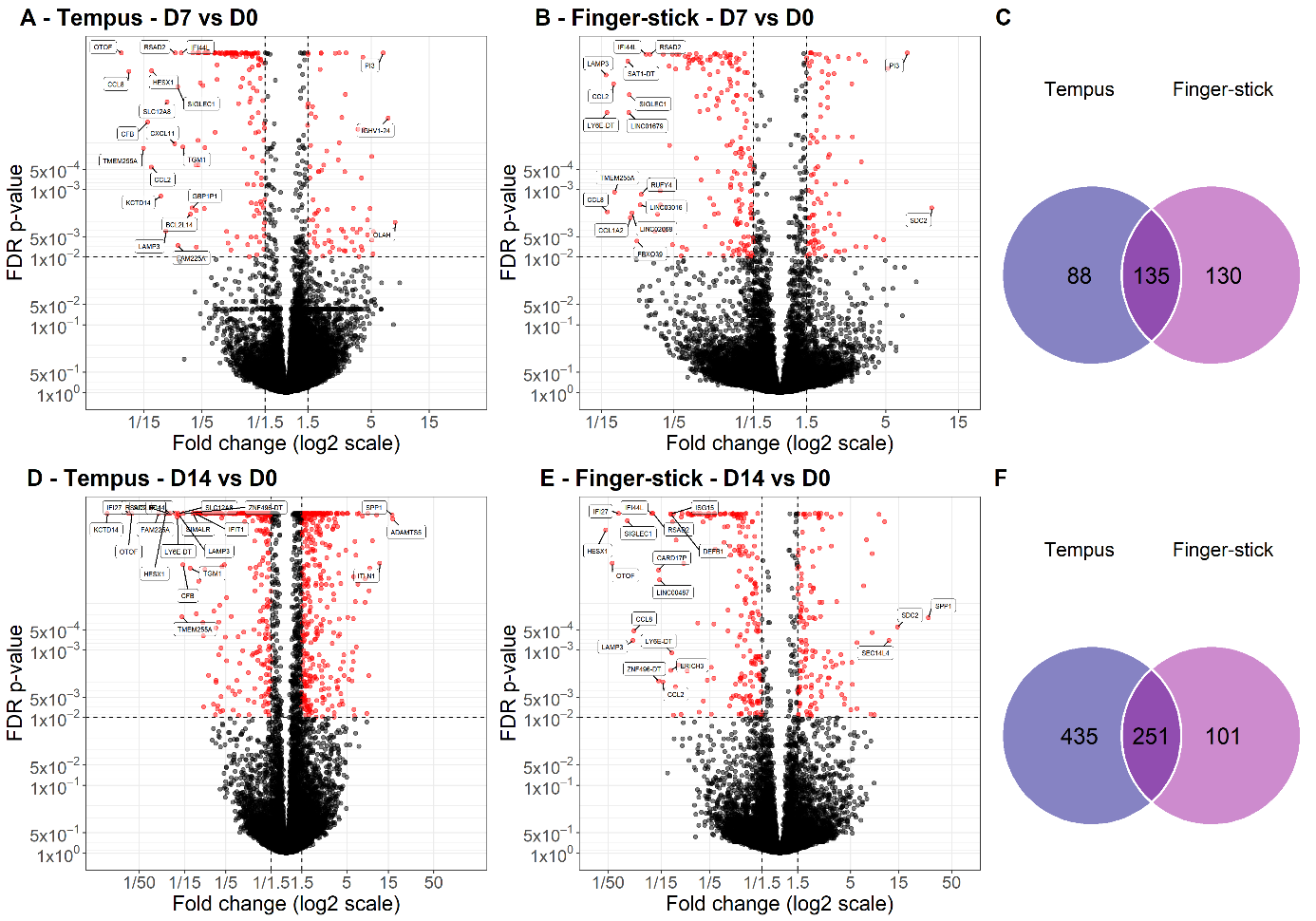

**Supplementary Figure 4. Differential abundance of genes at D7 and D14 compared to D0 in the 22 COVID19+ outpatient of the COVERAGE immune study. Blood recovered at D7 by Tempus (A) and finger-stick (B). Blood recovered at D14 by Tempus (D) and finger-prick (E).**

**The volcano plot displayed the top 20 differentially abundant genes. Venn diagram for the number of differentially abundant genes according to the sampling method at D7 (C) and D14 (F) both compared to inclusion (D0).**

Differential abundant genes with absolute FC > 1.5 and FDR < 0.01.

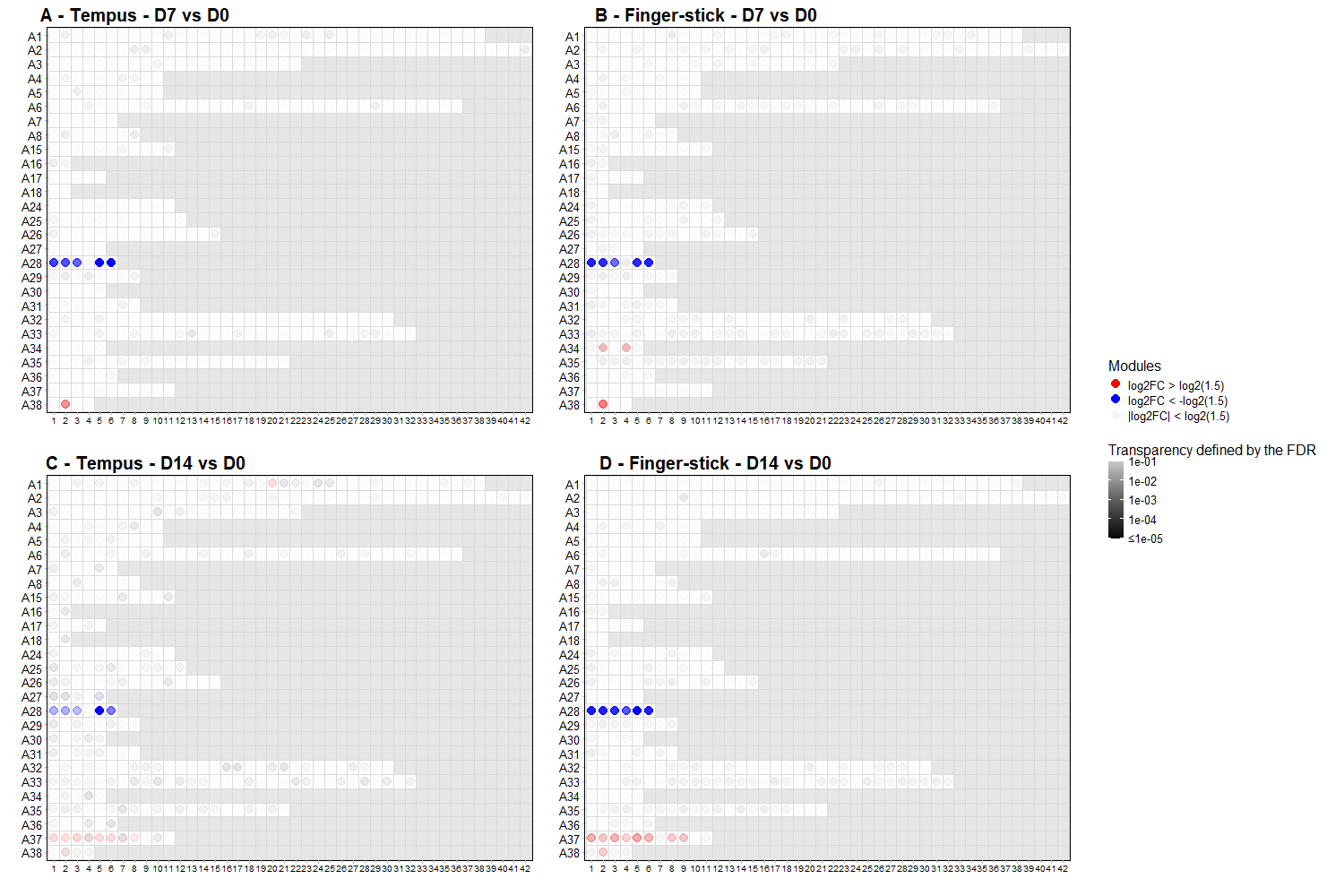

**Supplementary Figure 5.** **Description of the enriched gene sets measured in finger-stick (B, D) and venous (A, C) blood samples at D7 (A, B) and D4 (C, D), respectively, in the 22 COVID19+ outpatient of the COVERAGE immuno study.**

**A**

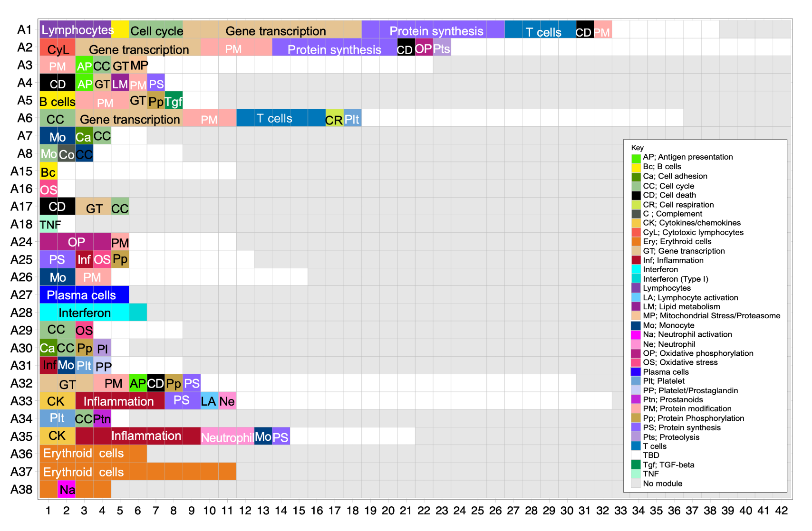

**B**

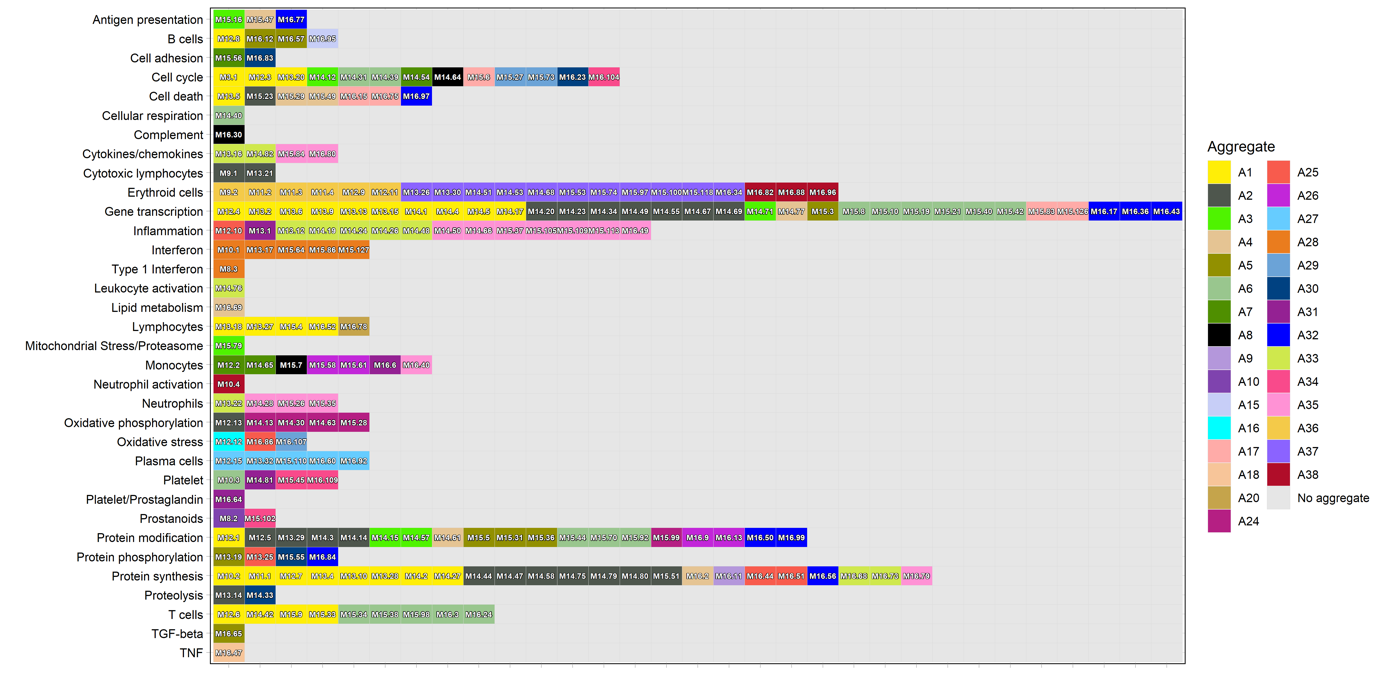

**Supplementary Figure 6. Reference grids (A, B) permit an overview of the aggregates and functions assigned to specific modules represented on the grid.**

Modules that are colored presented a differential abundance compared to baseline with absolute FC > 1·5 and FDR < 0·05.

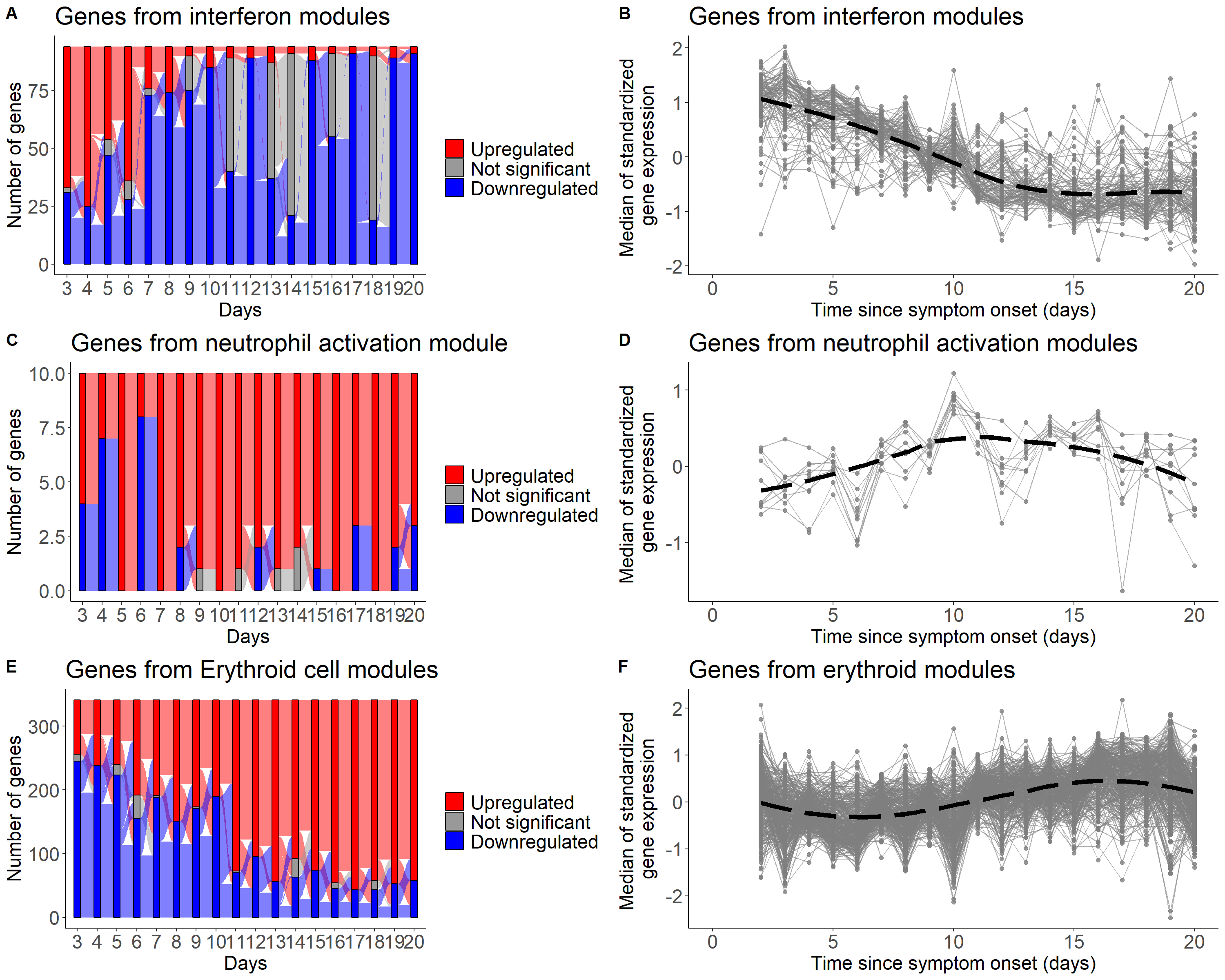

**Supplementary Figure 7. Change of gene abundance every days associated to interferon signaling, neutrophil activation and erythroid cells using finger-stick sampling**

Genes differentially expressed and evolution of abundance of genes of interferon (A-B), neutrophil activation (C-D) and erythroid (E-F) modules over time from symptom onset (each time point compared to inclusion)

**Supplementary Note 1**

In their reanalysis of data from the vaccine study by Obermoser *et al.*^1^, Stein *et al.*^2^ concluded that there were significant differences in gene expression between veinous and capillary (from finger-prick) blood. Unfortunately, we show here on the contrary that those data cannot be used to compare the two blood sources conclusively. The two factors preventing a truthful comparison are i) first the presence of an inextricable and strong batch effect confounded with the blood source sampling; and second ii) a sample size too small to allow for stratified analysis and further comparison of biological conclusions within each blood source.

Figure SN1.1 highlights the confusion between the flow-cells and the blood sources, while Figure SN1.2A shows the strong flow-cell batch effect. We used ComBat^3^ to correct this flow-cell batch effect — of note, we were unable to perfectly reproduce the results from Stein *et al.*^2^ as they provide only scarce information about their normalization pipeline of the microarray data; so, we relied instead on a state-of-the-art pipeline for pre-processing microarray data leveraging norm-exp background correction using negative control probes and quantile normalization using negative and positive control probes for normalization, as implemented in the limma R package. Figure SN1.2B highlights the impact of adjusting on blood source or not during the batch correction, demonstrating the confusion between this flow-cell batch effect and a potential difference between the two investigated blood sources.

Instead of using well-established multiple correction procedure — such as the Benjamini-Hochberg^5^ correction (which control the False Discovery Rate) — Stein *et al.*^2^ chose to use a 0.001 threshold on raw p-values for significance. This threshold, akin to using a Bonferroni correction when performing 50 tests, is nowhere near adequately accounting for the multiplicity of tests actually performed in this analysis, which compares gene expression between the two blood sources across 14,365 probes (after filtering out probes detected in less than 65% of the samples). Stein *et al.* ^2^ performed two different analyses using the paired moderated t-test from limma^6^: at each baseline time points (D-7 and D0) they tested for probes differentially expressed between blood sources (n=16 participants, 6 from each vaccine arm plus 4 placebo); in each of the two vaccine arms (Pneumo and Flu) they tested for probes differentially expressed at H12 or at H24 compared to baseline D0 within each blood source (n=6). Table SN1 summarize the number of significant probes identified for each of those pairwise comparisons with a fold-change greater than 1.25 according to the preprocessing of the data. We focus on three different preprocessing, either the lumi-normalized data without batch correction, or the limma-normalized data with correction for the flow-cell batch effect adjusted — or not — on the blood source. Interestingly, when correcting for the flow-cell batch effect, even while adjusting on the blood source, all differences vanish in the stratified analysis. This highlights the lack of power with only 6 individuals, especially compared to previous results using the larger sample size available from veinous blood^1,7^. At baseline, the probes identified with the adjustment further underline the inextricable confusion between the flow-cell batch effect and the potential blood source differences.

A

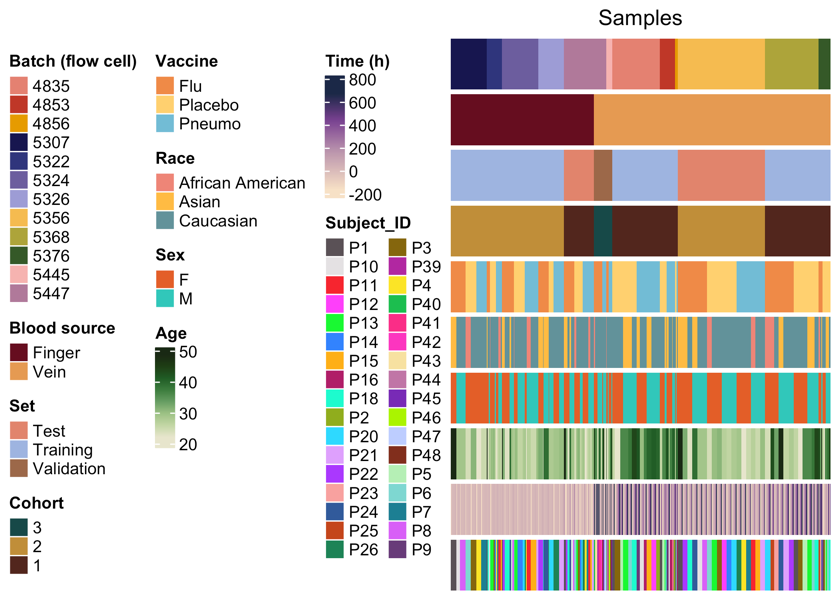

B

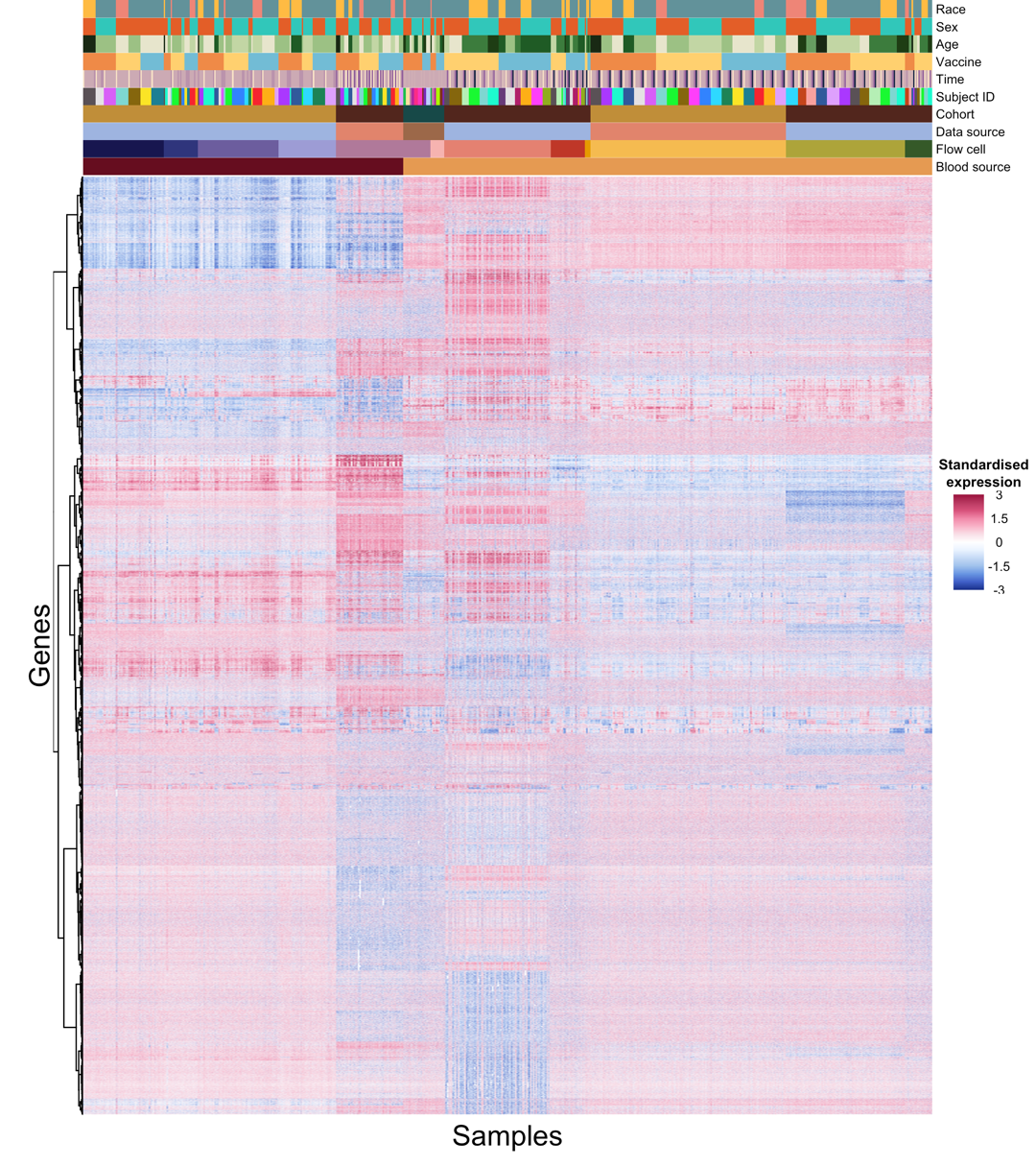

**Figure SN1.1: A.** Heatmap of the various metadata providing biological and technical information about the 621 microarrays available from Obermoser *et al.*^1^ highlighting the nesting confusion between flow-cells and blood source. **B**. Heatmap of the scaled gene expression from the same 621 samples ordered according to their respective blood source and flow-cell.

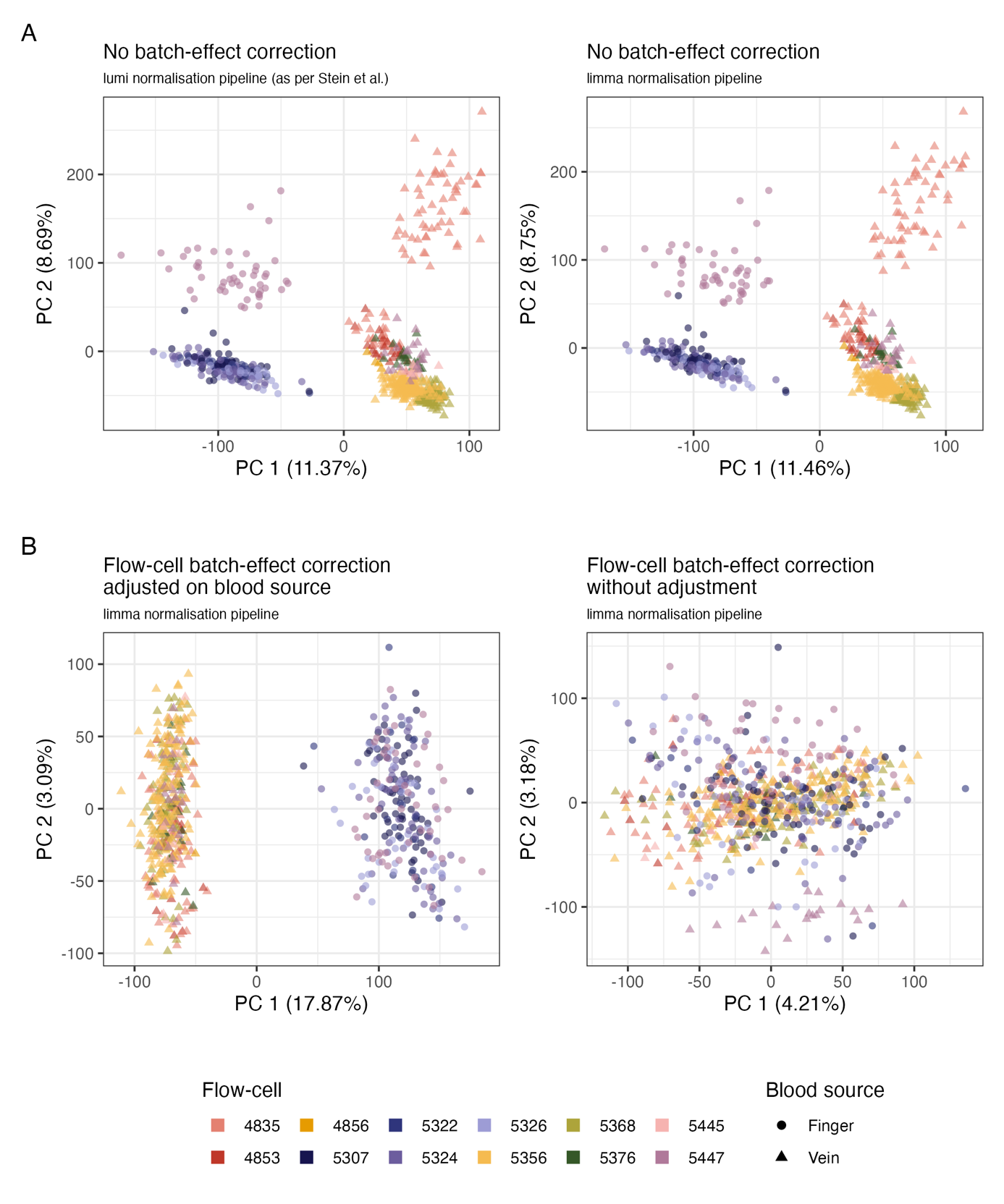

**Figure SN1.2:** First factorial plans of a Principal Component Analysis on four different pre-processing of the data. **A. left** lumi normalization pipeline without batch correction; **right** limma normalization pipeline without batch correction. Both normalization seem similar. **B. left** limma normalization pipeline with flow-cell batch correction adjusted on blood source; **right** limma normalization pipeline with flow-cell batch correction without adjustment.

| **Normalization** | **Batch correction** | **BC adjustment** | **Vaccine** | **Time** | **Blood source** | **Significant probes** |
| --- | --- | --- | --- | --- | --- | --- |
| lumi (as per Stein et al.) | no | - | baseline (n=16) | D-7 | both | 6480 |
| lumi (as per Stein et al.) | no | - | baseline (n=16) | D0 | both | 6416 |
| limma | yes | No | baseline (n=16) | D-7 | both | 0 |
| limma | yes | No | baseline (n=16) | D0 | both | 0 |
| limma | yes | on blood source | baseline (n=16) | D-7 | both | 740 |
| limma | yes | on blood source | baseline (n=16) | D0 | both | 685 |
| lumi (as per Stein et al.) | no | - | pneumo (n=6) | H12 | capillary | 168 |
| lumi (as per Stein et al.) | no | - | pneumo (n=6) | H12 | veinous | 82 |
| lumi (as per Stein et al.) | no | - | pneumo (n=6) | H24 | capillary | 1 |
| lumi (as per Stein et al.) | no | - | pneumo (n=6) | H24 | veinous | 0 |
| lumi (as per Stein et al.) | no | - | flu (n=6) | H12 | capillary | 0 |
| lumi (as per Stein et al.) | no | - | flu (n=6) | H12 | veinous | 0 |
| lumi (as per Stein et al.) | no | - | flu (n=6) | H24 | capillary | 0 |
| lumi (as per Stein et al.) | no | - | flu (n=6) | H24 | veinous | 0 |
| limma | yes | No | pneumo (n=6) | H12 | capillary | 0 |
| limma | yes | No | pneumo (n=6) | H12 | veinous | 0 |
| limma | yes | No | pneumo (n=6) | H24 | capillary | 0 |
| limma | yes | No | pneumo (n=6) | H24 | veinous | 0 |
| limma | yes | No | flu (n=6) | H12 | capillary | 0 |
| limma | yes | No | flu (n=6) | H12 | veinous | 0 |
| limma | yes | No | flu (n=6) | H24 | capillary | 0 |
| limma | yes | No | flu (n=6) | H24 | veinous | 0 |
| limma | yes | on blood source | pneumo (n=6) | H12 | capillary | 0 |
| limma | yes | on blood source | pneumo (n=6) | H12 | veinous | 0 |
| limma | yes | on blood source | pneumo (n=6) | H24 | capillary | 0 |
| limma | yes | on blood source | pneumo (n=6) | H24 | veinous | 0 |
| limma | yes | on blood source | flu (n=6) | H12 | capillary | 0 |
| limma | yes | on blood source | flu (n=6) | H12 | veinous | 0 |
| limma | yes | on blood source | flu (n=6) | H24 | capillary | 0 |
| limma | yes | on blood source | flu (n=6) | H24 | veinous | 0 |

**Table SN1.1:** Number of significant probes (FDR < 5% and absolute fold-change > 1.25) from paired moderated t-tests according to the data pre-processing.

In conclusion, our re-analysis comparing veinous and capillary blood from the Obermoser *et al*. study ^1^ show that i) confusion between a documented technical effect and the blood source disqualify this dataset from being able to provide meaningful insight about this comparison ii) using state-of-the-art and validated statistical tools for both pre-processing of the data and post-processing of the results drastically reduces the hypothetical differences originally put forth by Stein *et al.*^2^, casting further doubts on their actual existence in that study.
